# COVID-19 vaccination recruits and matures cross-reactive antibodies to conserved epitopes in endemic coronavirus Spike proteins

**DOI:** 10.1101/2022.01.24.22269542

**Authors:** Evan A Elko, Georgia A Nelson, Heather L Mead, Erin J Kelley, Virginia Le Verche, Angelo A Cardoso, Jennifer L Ely, Annalee S Boyle, Alejandra Piña, Sierra N Henson, Fatima Rahee, Paul S Keim, Kimberly R Celona, Jinhee Yi, Erik W Settles, George C Yu, Sheldon R Morris, John A Zaia, Jason T Ladner, John A Altin

## Abstract

The COVID-19 pandemic has triggered the first widespread vaccination campaign against a coronavirus. Most vaccinated subjects are naïve to SARS-CoV-2, however almost all have previously encountered other coronaviruses (CoVs) and the role of this immunity in shaping the vaccine response remains uncharacterized. Here we use longitudinal samples and highly-multiplexed serology to identify mRNA-1273 vaccine-induced antibody responses against a range of CoV Spike epitopes and in both phylogenetically conserved and non-conserved regions. Whereas reactivity to SARS-CoV-2 epitopes showed a delayed but progressive increase following vaccination, we observed distinct kinetics for the endemic CoV homologs at two conserved sites in Spike S2: these became detectable sooner, and decayed at later timepoints. Using homolog-specific depletion and alanine-substitution experiments, we show that these distinctly-evolving specificities result from cross-reactive antibodies as they mature against rare, polymorphic residues within these epitopes. Our results reveal mechanisms for the formation of antibodies with broad reactivity against CoVs.

## Introduction

Prior to the COVID-19 pandemic, widespread human immunity to viruses of the *Coronaviridae* family was limited to 4 species: the alphacoronaviruses HCoV-229E and HCoV-NL63, and the betacoronaviruses HCoV-OC43 and HCoV-HKU1. Now, global immunization to a fifth species – SARS-CoV-2 – is underway, through widespread natural infection, and increasingly through vaccination with formulations designed to immunize against the Spike protein, sometimes alongside other viral proteins. To date, the most effective and widely-studied vaccines contain modified mRNA encoding a stabilized full-length SARS-CoV-2 Spike protein (such as mRNA-1273 (Baden et al., 2021)), which generate robust T and B cell responses, including high titers of neutralizing antibodies that correlate with protection against disease (Bergwerk et al., 2021). It remains incompletely understood how pre-pandemic immunity to coronaviruses shapes this newly-acquired immunity against SARS-CoV-2.

One of the hallmarks of the polyclonal adaptive immune response is the generation of memory against a variety of epitopes of an infecting virus, sometimes including reactivity to conserved regions that can be recruited and matured in subsequent responses triggered by infections with heterologous viruses (White, 2021; Wong et al., 2020). Cross-reactive antibodies and memory B cells of this type support accelerated responses, but can have divergent functional consequences, ranging from life-threatening antibody-dependent enhancement (exemplified by dengue virus ((Halstead and O’Rourke, 1977)), to ‘imprinting’ that may constrain the specificity of subsequent responses (exemplified by influenza virus (Davenport et al., 1953; Gostic et al., 2016; Halstead and O’Rourke, 1977), and the maturation of antibodies with broad neutralizing capacity (exemplified by HIV ((Liao et al., 2013)).

The S1 subunit of the SARS-CoV-2 Spike protein diverges considerably in sequence from the corresponding regions of the four endemic seasonal coronaviruses, allowing the pandemic virus to evade most neutralizing antibodies raised by pre-pandemic coronavirus exposures (Poston et al., 2021). In contrast, the S2 subunit, containing the fusion apparatus, is more conserved and contains regions of high sequence identity to the other circulating betacoronaviruses, and in some cases also to members of the alphacoronavirus genus. Consistent with this conservation, antibody responses that cross-recognize Spike proteins from pandemic and endemic CoVs have been reported (Aguilar-Bretones et al., 2021; Anderson et al., 2021), and in some cases mapped to defined epitopes within S2 (Ladner et al., 2021; Pinto et al., 2021; Shrock et al., 2020; Song et al., 2021). These cross-reactive epitopes include the Fusion Peptide (FP) region, whose sequence is conserved across SARS-CoV-2 and the 4 endemic HCoVs, and the Stem-Helix (SH) region, conserved across many of the betacoronaviruses and involved in the conformational rearrangement that mediates membrane fusion. At the SH epitope, clones with the ability to broadly-neutralize across the betacoronavirus genus have been isolated (Pinto et al., 2021), however antibodies like these appear to comprise only a small fraction of the overall neutralizing response to SARS-CoV-2 Spike (Piccoli et al., 2020). A contribution to SARS-CoV-2 neutralization by antibodies binding the FP epitope has also been reported (Poh et al., 2020).

Here, we study the interaction between immunity to endemic CoVs and SARS-CoV-2, using longitudinal epitope-resolved profiling of plasma antibodies prior to and post-vaccination with the mRNA-1273 vaccine. PepSeq technology allows large programmable libraries of DNA-barcoded peptide constructs to be synthesized and assayed in massively-parallel reactions (Kozlov et al., 2012; Ladner et al., 2021). We use this platform to generate a novel human virome-wide 15,000-plex library of 30mer peptides representing the four HCoVs and SARS-CoV/SARS-CoV-2, map cross-reactive and non-cross-reactive Spike epitopes targeted by COVID-19 vaccination and study the evolution of the corresponding antibody specificities over time.

## Results

### Epitope-resolved antibody responses to CoV Spike proteins induced by vaccination

A cohort of 21 subjects with no prior SARS-CoV-2 infection history and undetectable SARS-CoV-2 antibodies were recruited prior to receiving both doses of the mRNA-1273 vaccine. Blood was drawn from each subject at baseline, and subsequently at post-vaccination days ∼8 (range: 7-10), ∼28 (range: 24-42, preceding the second dose) and ∼140 (range: 133-166) (**Figure 1a** and **Supplemental Table 1**). From a negative baseline, all subjects showed increases in total Spike-binding and Spike:receptor-blocking antibodies over time, beginning as early as day 8 in some cases, and uniformly progressing to strong positivity by the final timepoint (**Supplemental Figure 1**). To resolve the kinetics of antibody responses against conserved and non-conserved Spike epitopes, we analyzed plasma samples using the PepSeq platform (Ladner et al., 2021), in which plasma was incubated with a library containing 1000s of DNA-barcoded peptides, immunoprecipitated onto protein G beads, and then analyzed through bulk amplification and high-throughput sequencing of the DNA tags to identify the IgG-binding peptides.

**Figure 1:**
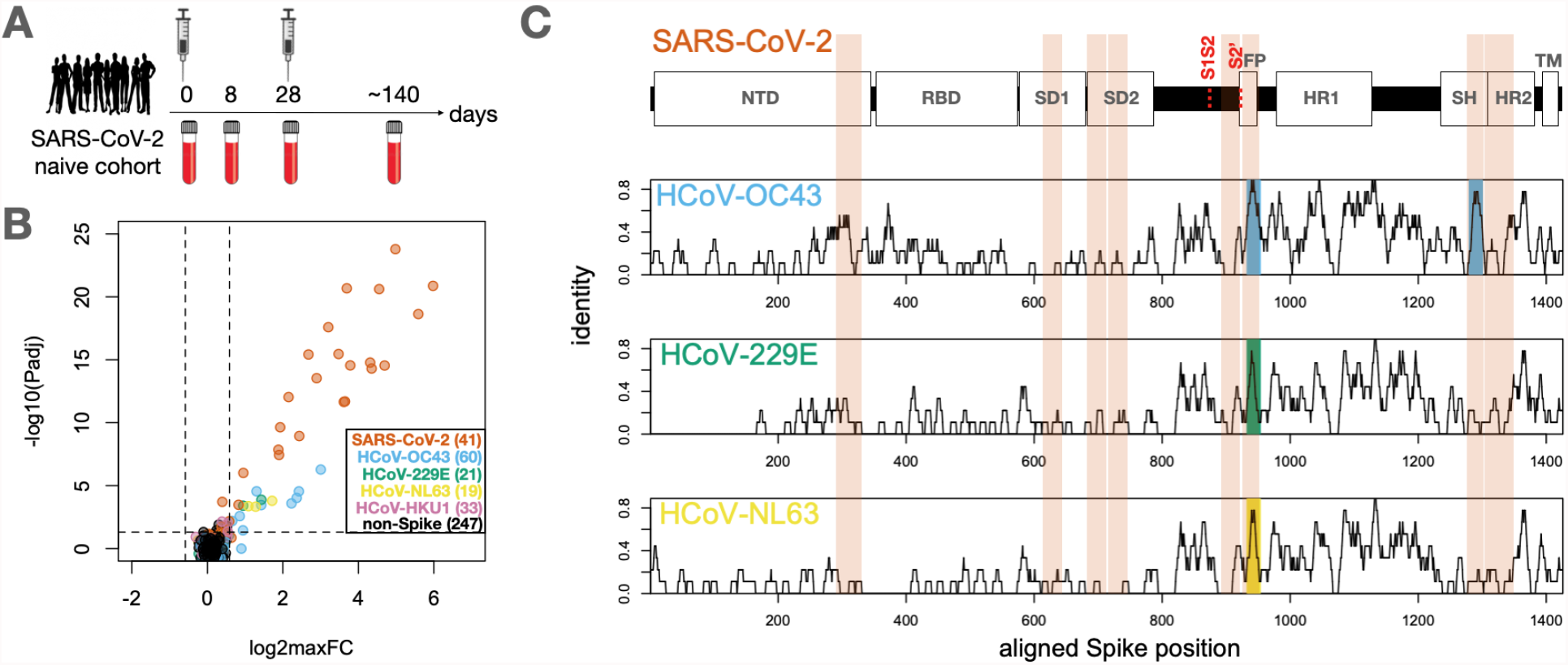
Identification of conserved and non-conserved antibody epitopes recognized following COVID-19 vaccination. **A**. A cohort of 21 subjects with no prior SARS-CoV-2 infection history and undetectable SARS-CoV-2 antibodies prior to vaccination received two doses of the mRNA-1273 vaccine, and gave blood samples at days 0, ∼8, ∼28 and ∼140 days relative to the first dose. **B**. Plasma from each timepoint was analyzed by PepSeq with a 15,000-peptide human virome library (‘HV2’). Z-scores for the 421 HV2 peptides designed from members of the *Coronaviridae* family were analyzed across subjects and timepoints to identify peptides showing significant time-differential signal. Each dot represents an individual peptide: Y-axis shows the FDR-adjusted ANOVA P-value (Padj) across all timepoints, and X-axis shows maximum log2 fold change (log2maxFC), which was calculated by dividing maximum Z-score at day 8, 28, and 140 by the Z-score at day 0. Legend in the lower-right shows the number of peptides for each CoV Species. Dotted lines indicate thresholds at Padj=0.05 and maxFC=1.5. **C**. Mapping of differentially recognized peptides (passing both Padj and maxFC thresholds shown in B) to a multiple sequence alignment of Spike proteins from SARS-CoV-2, HCoV-OC43, HCoV-229E, and HCoV-NL63. Plots show structural features or cleavage sites (dotted red lines) of SARS-CoV-2 Spike (*upper*) and sliding-9mer window amino acid sequence identity to SARS-CoV-2 Spike for the three endemic Spike proteins (*lower*). The differentially-recognized peptides map to 8 epitopes in SARS-CoV-2 spike (vertical orange boxes), 2 epitopes in HCoV-OC43 (cyan), 1 epitope in HCoV-229E (gold), and 1 epitope in HCoV-NL63 (green). Epitopes detected in the three endemic CoV proteins all occur at two regions of high sequence conservation with SARS-CoV-2. The following features of Spike are highlighted: NTD=N-terminal Domain; RBD=Receptor Binding Domain; SD1=Subdomain 1; SD2=Subdomain 2; FP=Fusion Peptide; HR1=Heptad Repeat 1; SH=Stem Helix; HR2=Heptad Repeat 2; TM=Transmembrane region.

For these studies, we designed and synthesized a PepSeq library (‘HV2’) containing 15,000 30mer peptides representing 80 human-infecting viral species. These sequences were each selected based on some *a priori* evidence of reactivity, including (i) an empirical PepSeq screen conducted on a larger library of peptides, (ii) published epitopes reported in the Immune Epitope Database, and (iii) homology to known reactive epitopes in related species (see **Methods** for details). Within the HV2 library, 421 peptides were designed from five Coronaviridae (CoV) species (SARSr-CoV, HCoV-OC43, HCoV-229E, HCoV-NL63 and HCoV-HKU1), of which 174 were derived from Spike sequences. An additional 60 sequences were included in which two 30mer peptides containing SARS-CoV-2 Spike epitopes were subjected to saturation Alanine scanning (discussed further below).

To identify peptides recognized by vaccine-responsive antibodies, we used ANOVA to compare the responses to each of the CoV peptides over time in each subject. Vaccine-responsive peptides were defined as those with FDR-corrected p-value <0.05 and average fold-induction across the cohort (maximum post-vaccine value relative to day 0) > 1.5. As expected, none of the 247 non-Spike-derived peptides met these criteria (**Figure 1b**). In contrast, 22 of the 41 SARS-CoV-2 Spike peptides were strongly responsive (**Figure 1b**), with average fold-inductions across the cohort ranging up to ∼64. In addition, 11 Spike peptides from other CoVs (7 from HCoV-OC43, 3 from HCoV-NL63, and 2 from HCoV-229E) were significantly vaccine-responsive, but more weakly so, with fold indications ranging up to ∼8.

Mapping of vaccine-responsive peptides to a multiple sequence alignment of the four Spike proteins revealed eight epitope regions (**Figure 1c:vertical orange boxes**): 4 in the S1 subunit, 3 in the S2 subunit and 1 in the S1S2 region, several of which have been previously described as targets of anti-SARS-CoV-2 reactivity (Ladner et al., 2021; Shrock et al., 2020). SARS-CoV-2 peptides were reactive at all 8 of these epitopes. In contrast, reactivity to the three endemic HCoVs was restricted to two regions in the S2 subunit: the Fusion Peptide (FP) and Stem Helix (SH) regions (**Figure 1c: cyan, gold, green shading**). In each case, the pattern of reactivity detected by PepSeq reflects epitope sequence conservation with SARS-CoV-2 (**Figure 1c: black traces**). Specifically, the FP region which is highly conserved across both the alpha and betacoronavirus genera (which include all human-infecting CoVs), was targeted by the vaccine response in all four of the aligned Spike proteins. In contrast, the SH region, which is conserved across only the betacoronavirus genus (including SARS-CoV-2 and HCoV-OC43, but not HCoV-NL63 or HCoV-229E), was recognized by vaccine-induced antibodies targeting only the respective betacoronavirus-derived sequences.

### Differential kinetics of the vaccine response across epitope classes

Next, we studied the kinetics of the vaccine-induced responses across the identified epitopes. Vaccine-responsive peptides were organized into the 8 epitopes described above (6 non-conserved and 2 conserved – sequences for each are listed in **Supplemental Table 3**) and the maximum signal for each was calculated at each timepoint in each subject. Time-resolved reactivity patterns partitioned into 2 groups: a “late-progressive” pattern for SARS-CoV-2 epitopes and an “early peak” pattern for endemic epitopes (**Figure 2a,b**). Specifically, reactivity to each of the 8 SARS-CoV-2 epitopes was not significantly increased at day 8, intermediately and variably increased at day 28, and universally and maximally increased at day 140 (**Figure 2b**). In contrast, reactivity to endemic homologs at the FP and SH epitopes was elevated as early as day 8, maximal at day 28 and then declined from day 28 to day 140 (**Figure 2b**).

**Figure 2:**
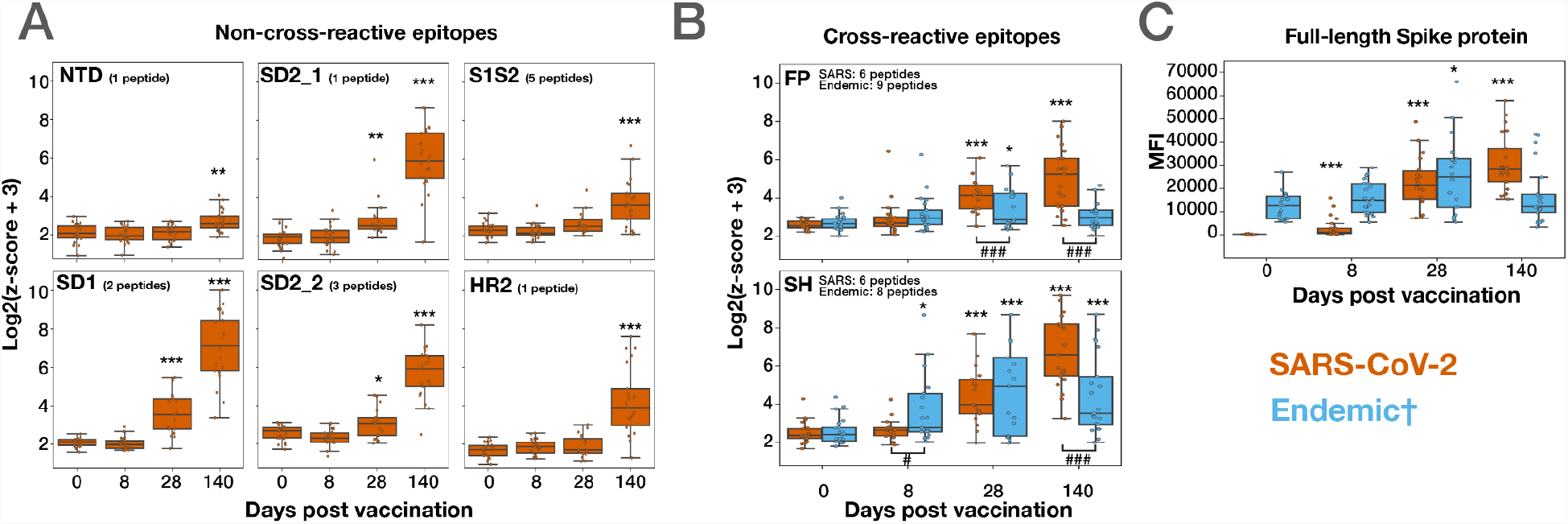
Divergent kinetics of the responses to SARS-CoV-2 compared to endemic Spike antigens following vaccination. PepSeq Z-scores for the six non-cross-reactive epitopes (**A**) and two cross-reactive epitopes (**B**) across the vaccinated cohort at timepoints of approximately 0, 8, 28 and 140 days post-vaccination. Shown is the maximum Z-score from the collection of peptides overlapping each vaccine-responsive epitope (Figure 1). The title of each plot indicates the focal epitope, named according to the features shown in Figure 1C. **C**. Antibody reactivity against full-length SARS-CoV-2 and HCoV-OC43 Spike proteins in vaccinated subjects across time, detected using an fluorescent bead assay. Across all panels, orange boxes indicate the response to the respective SARS-CoV-2 peptides/protein, whereas blue boxes indicate reactivity to the respective endemic peptides/protein. † The “Endemic” category includes peptides from HCoV-OC43, HCoV-229E and HCoV-NL63 for the FP epitope in **B**, peptides from HCoV-OC43 for the SH epitope in **B**, and full-length HCoV-OC43 Spike protein in **C**. The limits of the boxes correspond to the 1st and 3rd quartiles, the black lines inside each box correspond to the median and the whiskers extend to points that lie within 1.5 interquartile ranges of the 1st and 3rd quartiles. * represents comparisons between the indicated time point and Day 0. t test:*p < 0.05,**p < 0.01,***p < 0.001. # represents comparisons between SARS-CoV-2 and Endemic epitopes within individual timepoints. t test ### p<0.001.

To determine how these epitope-level kinetics compared to the anti-Spike antibody response more generally, we analyzed the same samples using multiplexed assays in which plasma IgG cross reactivity to full-length SARS-CoV-2 and HCoV-OC43 Spike proteins was quantified using target-conjugated fluorescent magnetic beads (MagPix, Luminex Corp.). Similar to the results of the epitope-resolved assay, we observed significant vaccine-induced antibody responses to both targets, but with different kinetics. Whereas the response to SARS-CoV-2 Spike increased from day 0 to day 140, the response to the endemic betacoronavirus OC43 was elevated at day 28, but decreased back to baseline by day 140 (**Figure 2c**). Therefore, we conclude that the response kinetics for individual peptide epitopes detected by PepSeq reflect those of the broader composite responses against the pandemic and endemic Spike proteins.

### Quantification of antibody cross-reactivity at the FP and SH epitopes over time

The induction of a response against conserved regions of endemic CoV Spike proteins following vaccination with SARS-CoV-2 Spike suggests the existence of antibodies capable of cross-recognizing both endemic and SARS-CoV-2 homologs at each of the FP and SH epitopes. To formally test this hypothesis, we performed cross-depletion experiments in which plasma was pre-treated with beads bearing pooled FP and SH peptides from either SARS-CoV-2 or HCoV-OC43 (hereafter the ‘depleting species’) to deplete species-specific subsets of antibodies, prior to being assayed by PepSeq. In this scheme, reactivity to peptides from the depleting species is expected to decrease (relative to undepleted samples), and any reduction in reactivity to the non-depleting species would imply the existence of antibodies capable of cross-recognizing both species (**Figure 3a**).

**Figure 3:**
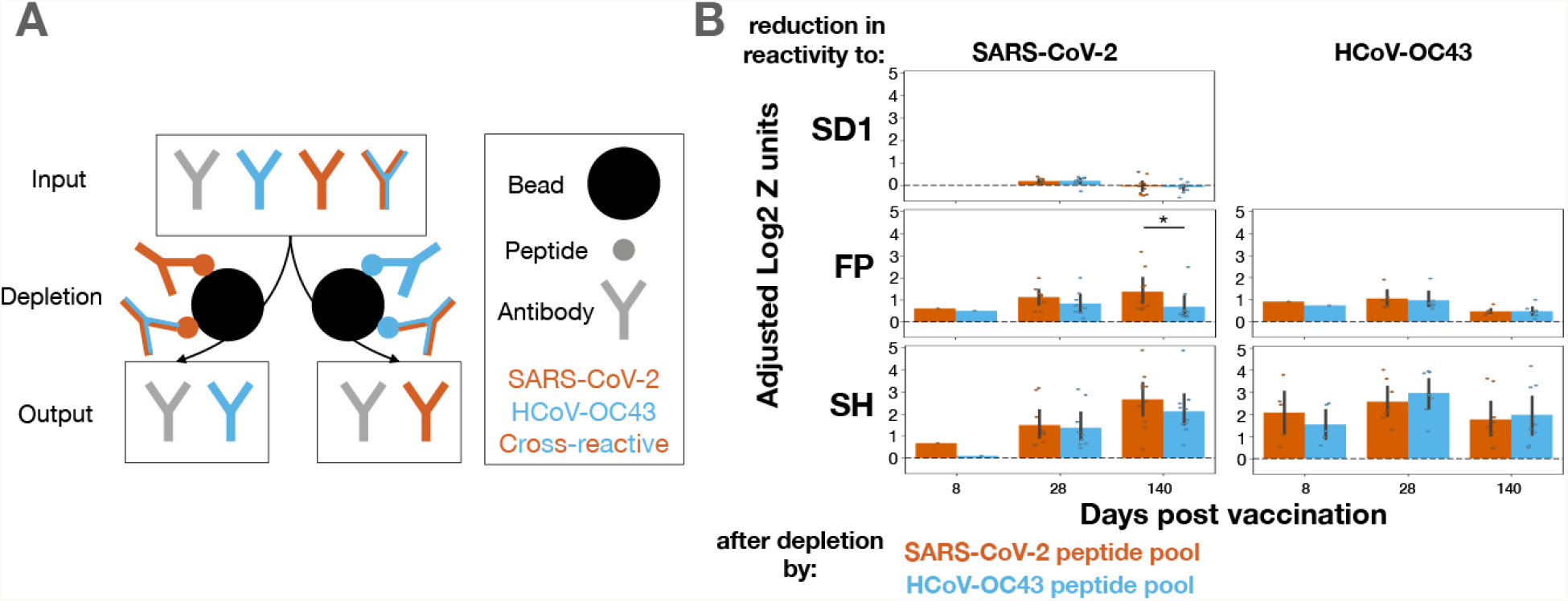
Cross-reactive antibodies dominate the responses to FP and SH across time, with the emergence of late mono-reactivity to SARS-CoV-2 FP. **A**. To quantify the extent of cross-reactivity between species at the two conserved Spike epitopes, vaccinnee plasma samples were treated with beads bearing biotinylated FP+SH peptide pools of either the SARS-CoV-2 (orange) or HCoV-OC43 (blue) sequences to deplete binding antibodies. The resulting samples were assayed by PepSeq, wherein a reduction in signal corresponding to the non-depleting species would indicate the presence of cross-reactive antibodies. **B**. Reduction in PepSeq signal for the SARS-CoV-2 and/or HCoV-OC43 homologs of the SD1 (SARS-CoV-2 only, upper), FP (middle) or SH (lower) epitopes at the indicated post-vaccination timepoint, after depletion by FP+SH peptide pools of either the SARS-CoV-2 (orange) or HCoV-OC43 (blue) sequences. Black bars show 95% confidence interval. Plots include only subject:timepoint combinations with detectable reactivity for either species at the focal epitope (Z-score > 6). t test:*p < 0.05,***p < 0.001

While we observed no change in signal for epitopes not included in the depleting pool (**Figure 3b - top row**), strong decreases were evident in the reactivity to both the SARS-CoV-2 and HCoV-OC43 homologs following depletion with FP and SH peptides from either species, revealing substantial cross-reactivity (**Figure 3b - middle and bottom rows**). Indeed, for the SH epitope, no difference was detectable at any timepoint in the extent of signal reduction for the depleting species compared to the non-depleting species, indicating that cross-reactive antibodies dominate the response. Similarly, signal reduction was unchanged between the depleting and non-depleting species for the FP epitope, except at the final timepoint where significantly less of the response to the SARS-CoV-2 epitope was depleted by the HCoV-OC43 peptide (**Figure 3b – middle panel**). We conclude that the responses against these two conserved epitopes are dominated across time by antibodies that cross-recognize both species, with the exception of the late response to FP, in which SARS-CoV-2 mono-reactivity becomes detectable.

### Trajectories of the maturing response at cross-reactive epitopes

To examine the temporal trajectories of antibody maturation in individual subjects, we derived a ‘maturation’ index for each of the cross-reactive epitopes, which represents the ratio of reactivity to the SARS-CoV-2 versus endemic peptides at each epitope. To avoid noise, we restricted this analysis to cases in which there was some overall detectable reactivity at the region of interest by excluding those donor-timepoint combinations in which no peptide (from any species) at the focal epitope exceeded a Z-score threshold (**Methods**). Applying this criterion, we detected FP reactivity in 3, 8, 10 and 15 donors, and SH reactivity in 4, 4, 8, 15 donors, respectively, at days 0, 8, 28 and 140 (**Figure 4a**). Although this metric of maturation varied widely across donors at each timepoint, there was a significant increase over time for both the FP and SH epitopes. More specifically, when maturation trajectories were resolved to the level of individual donors, we observed that responses began at a range of maturation levels, but then consistently underwent progressive increases over time, beginning at day ∼8.

**Figure 4:**
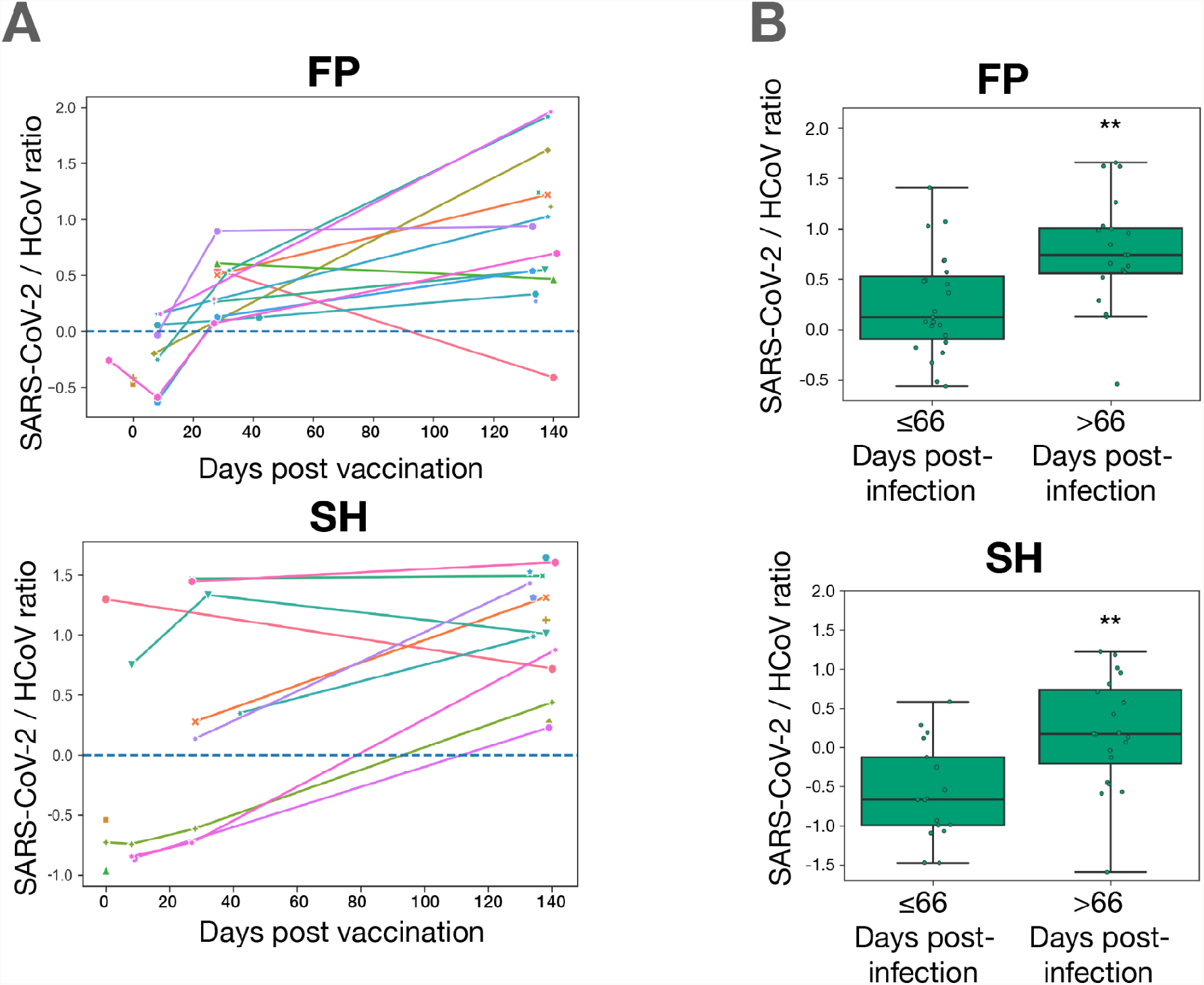
Temporal maturation of antibody specificity at the FP and SH epitopes in ≥vaccinated and convalescent subjects. **A**. Log-transformed ratio of PepSeq Z-scores for the SARS-CoV-2 and endemic HCoV homologs of the FP and SH epitopes plotted against days post-vaccination. Shown are data points in which reactivity against the focal epitope exceeds the threshold of detection(FP:Z-score ≥ 4,SH:Z-score ≥ 10) (in either the SARS-CoV-2 or HCoV homologs), and lines connect data points from the same subjects. **B**. Maturation of the antibody responses at the FP and SH epitopes in unvaccinated COVID-19 convalescent subjects dichotomized by days post-infection and analyzed using the ratio described in A. t test:**p < 0.01

To determine whether a similar maturation of the antibody response at these two cross-reactive epitopes also occurs in the setting of SARS-CoV-2 infection, we studied a cohort of 46 COVID-19 convalescent subjects from whom plasma was collected at a range of timepoints following infection (range: 17-142 days, one sample per subject) (**Figure 4b** and **Supplemental Table 2**). These samples were assayed by PepSeq using the HV2 library, and analyzed for FP and SH maturation using the same metric as described for the vaccinated cohort. Donor-level longitudinal analysis was unavailable as the convalescent cohort lacked serial collections, however dichotomization of subjects according to the number of days between diagnosis and plasma collection revealed significant increases in maturity in the later cohort for both the FP and SH epitopes (p-value: 0.0023, 0.0018 respectively).

### Fine mapping of the Spike residues recognized by maturing, cross-reactive responses

To study the mechanisms by which antibodies may cross-recognize – yet also mature to distinguish – the SARS-CoV-2 and endemic homologs of the FP and SH epitopes, we used alanine-substitution scanning to map sequence dependencies of the vaccine-induced antibody responses at amino-acid resolution (**Figure 5**). For each position in both epitopes, we calculated a ‘residue dependency index’, representing the log2 ratio of signal from the wild-type peptide versus a mutant peptide with alanine substituted at the focal site. Analysis across Spike positions 810-839 (containing FP) and 1136-1165 (containing SH) in day 140 post-vaccination samples revealed core regions of dependency for each epitope (positions 814-825 for FP and 1148-1156 for SH) that were largely consistent across donors (**Figure 5a**). For each epitope, the core closely matched the region of maximum sequence identity across CoV species, consistent with the observed cross-reactivity. However, for both epitopes we also observed some dependence on positions that were polymorphic between viruses: positions K814 and I818 in the FP epitope were involved in binding for most donors, and in >90% of subjects the anti-SH response was heavily dependent on the Y1155 residue. Two polymorphic sites outside of the core binding region (D830 and I834) also contributed to antibody binding in most donors at the FP epitope. Overall, the contribution of polymorphic sites within otherwise conserved regions to antibody binding provides a mechanism by which the response may distinguish closely-related homologs at each of these cross-reactive epitope regions.

**Figure 5:**
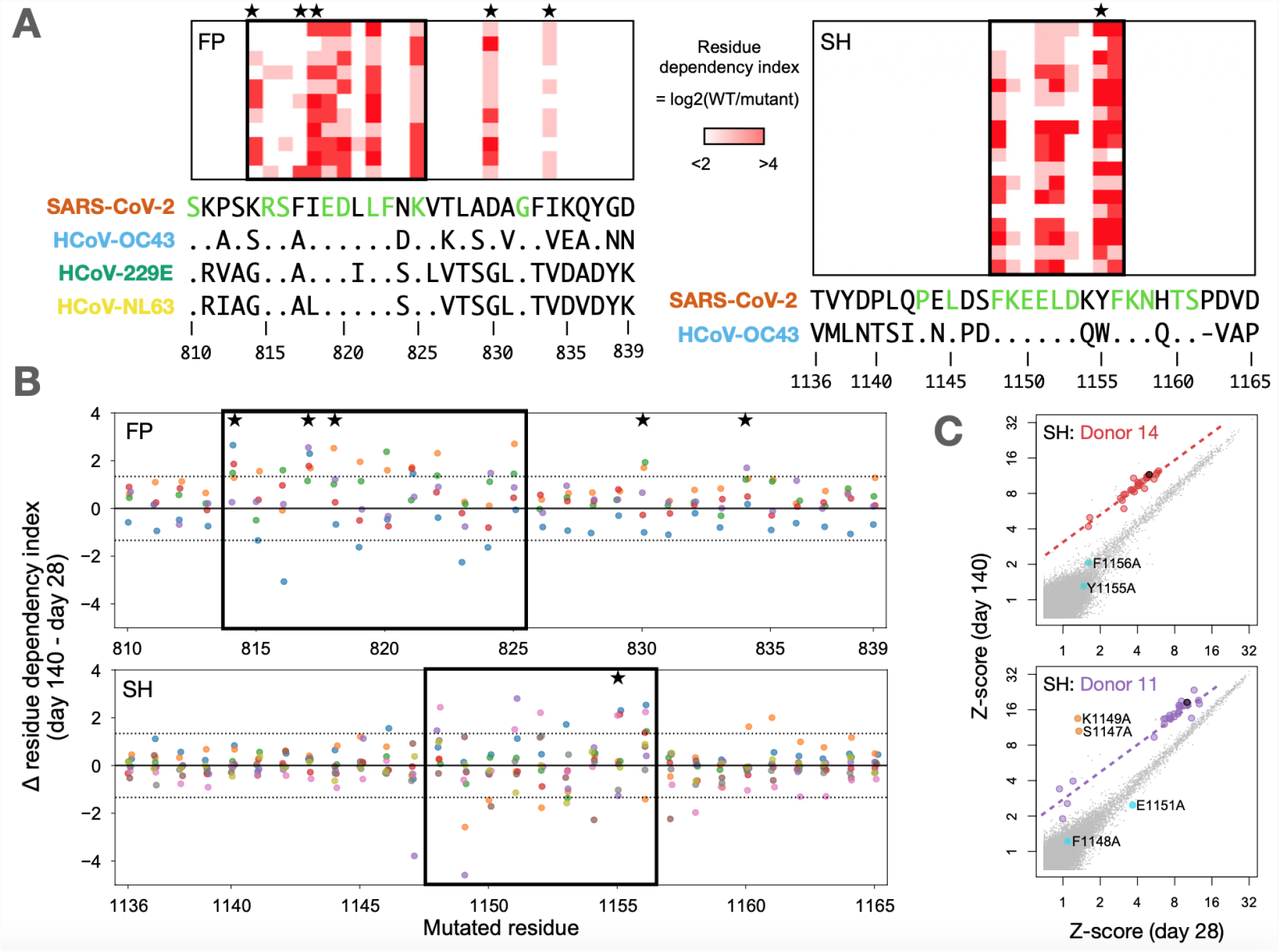
Matured responses to FP and SH epitopes depend on both conserved and polymorphic residues. **A**. PepSeq analysis of vaccinated subjects (rows) at day 140 for sets of HV2 peptides (columns) in which each residue of SARS-CoV-2 Spike at positions 810–839 (containing FP region, *left*) or positions 1136-1165 (containing SH region, *right*) was individually substituted to alanine. Color scale indicates the log2(ratio) of PepSeq Z-scores in each subject for the wild-type peptide, divided by the respective alanine-substituted peptide (‘residue dependency index’). Boxes enclose the inferred core binding regions, and the sequence letters colored in green below indicate positions that are conserved between all species shown. Stars indicate polymorphic positions that are involved in binding. Shown are subjects for whom the response to the respective wild-type peptide exceeds a Z-score threshold of 10: respectively, 11 and 18 subjects for FP and SH. **B**. Analysis of the Alanine-substituted FP (*upper*) and SH (*lower*) peptide sets described in A, now comparing the responses at day 28 v day 140 for subjects (each in a different color) whose reactivity to the wild-type peptides at both timepoints exceeded a Z-score threshold of 10: respectively, 5 and 9 subjects for FP and SH. The y-axis shows the log2 change (from the day 28 to the day 140 timepoint) in residue dependency (as defined in A), at the position shown on the x-axis. Dotted horizontal lines demarcate the 99th percentile range of variation observed in a set of 1830 control data points from the same 9 subjects at 11 non-CoV epitopes to which stable (non-vaccine-induced responses) were detected and that were also each represented as sets of 30 alanine mutants and analyzed in the same way. **C**. Example scatterplots showing peptide-level reactivity at day 28 v day 140 in 2 subjects with evidence for temporal changes in residue-level specificity within the SH epitope. Alanine-mutated SH peptides are highlighted in red/purple (corresponding to their respective donor’s color in B), the wild-type SH peptide in black, and non-SH peptides in gray. Peptides with significant temporal deviation relative to wild-type (as calculated in B) are shown in cyan (increasing dependency on the substituted position) or orange (decreasing dependency on the substituted position), and annotated with the coordinate of the substituted residue. Dotted lines show the fit for mutant peptides without significant temporal deviation.

To study the evolution of fine-specificity at the FP and SH epitopes, we compared residue dependencies between day 28 and day 140 for a subset of 5 and 9 subjects with robustly-detectable reactivity (Z ≥ 10) at both days (**Figure 5b, c**). Using a set of stably-recognized non-CoV viral epitopes that were also each represented in the form of 30 alanine mutants in the HV2 library, we set a threshold that bounded 99% of changes in residue dependency between day 28 and day 140 observed under conditions presumed to be non-maturing for those epitopes (horizontal dotted lines in **Figure 5b**; see **Methods**). In total, 29 and 28 events (of a total 150 and 270 subject / position combinations) were observed above this threshold for FP and SH respectively (p<1e-9), of which 25 (86%) and 22 (79%) occurred in the inferred core regions. In some cases, residues that were essential for binding at day 28 became largely dispensable at day 140 (eg purple dots below the midline at SH residues S1147, K1149 in **Figure 5b**), and conversely, we observed examples where positions with small contributions to binding at day 28 became substantially more important at day 140 (eg blue dots above the midline at SH residues D1153, Y1155 and F1156 in **Figure 5b**). For both epitopes, this latter category included polymorphic positions – eg K814 (3 donors), F817 (4 donors) and D830 (2 donors) in FP, and Y1155 (3 donors) in SH – indicating their increased involvement as the response matures. We conclude that maturation of the vaccine response to cross-reactive epitopes includes temporal changes in the fine-specificity of the response, including at positions that are polymorphic between CoV species.

## Discussion

Using longitudinal samples and highly-multiplexed serology, we analyzed the circulating antibody response following COVID-19 vaccination and its evolution over time at epitope-level resolution. We showed that vaccination elicits robust antibody responses against a range of Spike epitopes from SARS-CoV-2, as well as several Spike epitopes from other HCoVs, and that these responses partition into two classes. The first class is directed against non/minimally-conserved regions of the Spike protein and involves antibody responses that are undetectable until day 28 post-vaccination, but then strongly increase after the second vaccine dose. We did not observe antibody reactivity against homologous HCoV peptides for epitopes in this class. The second class of antibodies are directed against two sites in the S2 subunit – the Fusion Peptide (FP) and Stem Helix (SH) – that are highly-conserved with Spike proteins from endemic HCoVs. Whereas reactivity to the SARS-CoV-2 sequences at these two epitopes follows the same kinetics as the first class (ie undetectable until day 28, and then increasing through day 140), binding to the corresponding endemic homolog epitopes is observed as soon as day 8 post-vaccination and then decreases between day 28 and day 140 (**Figure 2**).

In general, the most parsimonious explanation for disparate kinetics among responses recognizing closely-conserved homologs is that they reflect the affinity maturation of cross-reactive antibodies. At both the FP and SH epitopes, our results provide several additional lines of direct evidence for antibodies capable of cross-recognizing both the SARS-CoV-2 and endemic peptide sequences. Most fundamental is our observation that responses against the endemic versions of both epitopes – as well as the full-length HCoV-OC43 Spike protein – are significantly induced following vaccination (**Figure 1**), indicating that antibodies stimulated by the SARS-CoV-2 immunogen also bind the endemic homologs, consistent with prior studies (Amanat et al., 2021). More specifically, in cross-depletion experiments focused on both the FP and SH epitopes (**Figure 3**) we observe that beads coated with peptides corresponding to either species generally deplete indistinguishable fractions of the antibody reactivity to the SARS-CoV-2 and endemic homologs. We observed this pattern at all post-vaccine timepoints for SH, and the two earliest timepoints for FP, indicating that the responses at these sites are generally dominated by cross-reactive antibodies. The exception to this rule is the response to SARS-CoV-2 FP at day 140, a substantial component of which is depletable by the SARS-CoV-2 sequence but not the endemic homolog. This indicates that a late mono-specific reactivity arises at this epitope, and could be explained by the acquisition of somatic mutations that reduce the endemic-specific binding of cross-reactive clones and/or the emergence of new clones specific for SARS-CoV-2 alone.

Consistent with the observed cross-reactivity, fine mapping at the FP and SH regions of Spike (**Figure 5**) confirms that the core antibody recognition sites reside in regions of high amino acid sequence identity – within much of the betacoronavirus genus in the case of SH, and spanning the alpha and betacoronavirus genera in the case of FP. Within each core recognition site, the SARS-CoV-2 epitopes differ by 2-3 amino acids from their endemic homologs, indicating residues whose differential participation in antibody binding can allow the response to distinguish between species. In support of this mechanism, our alanine-scanning results reveal several examples in which the response shows selectively increased dependence on non-conserved residues as it evolves from day 28 to day 140. The ability to focus the evolving specificity of recruited antibodies on a small minority of non-conserved residues is consistent with the sustained germinal center responses observed after infection (Dugan et al., 2021; Sokal et al., 2021) and vaccination (Turner et al., 2021) and underscores the remarkable discriminating power of such responses, which can serve as an important safeguard against the production of autoantibodies in cases of molecular mimicry between self and non-self (Burnett et al., 2018). Future studies tracking the sequences of individual cross-reactive antibody clones as their specificities evolve from endemic to SARS-CoV-2-dominant recognition, including in combination with structural analysis, will allow the molecular basis of the evolution between Spike S2 homologs to be resolved in further detail.

The functional consequences of the cross-reactive responses to Spike S2 remain to be characterized, however previous studies in convalescent patients (Poh et al., 2020); (Pinto et al., 2021; Sauer et al., 2021) and immunized animals (Ravichandran et al., 2021; Wang et al., 2021) have shown that antibodies against both the FP and SH epitopes have neutralizing activity against SARS-CoV-2. Exposure and incorporation of the fusion peptide region of Spike into the host cell membrane are essential steps in virus entry into cells, and the corresponding site in the HA2 subunit of influenza A Hemagglutinin is also conserved and has been shown to be targeted by antibodies with broad neutralizing activity (Corti et al., 2011). FP is also known to be the target of an immunodominant CD4 T cell response to SARS-CoV-2 that cross-reacts with endemic CoVs (Low et al., 2021; Loyal et al., 2021).

At the SH epitope, Pinto and colleagues describe monoclonal antibodies recognizing the precise epitope we identify here, and capable of neutralizing multiple members of the betacoronavirus genus, albeit with modest potency (Pinto et al., 2021; Sauer et al., 2021). Interestingly, however, they observe a prevalence of antibodies directed to this epitope (∼20% of vaccinees, ∼20% of convalescent subjects) that is substantially lower than the rate described here (>90% of vaccinees) and in previous work (∼50-80% of convalescent subjects: (Kaslow et al., 2014; Ladner et al., 2021; Shrock et al., 2020)). Together with evidence that antibodies against non-RBD targets generally account for <10% of the overall neutralizing activity in convalescent donors (Piccoli et al., 2020), these observations suggest that antibodies with measurable neutralizing function against multiple human-infecting CoV species may arise commonly following SARS-CoV-2 infection or vaccination, but play a limited role in overall neutralization. Possibly exacerbated by hindered antibody access to the Spike stalk (ref), a lack of widespread, strongly cross-protective anti-S2 responses (and the associated selective pressure for immune escape) would be consistent with the high conservation of this region across human-infecting CoV species.

Our work suggests that, in addition to protection against SARS-CoV-2, COVID-19 vaccination may induce a boost in immunity against seasonal endemic CoVs, mediated by cross-reactive epitopes such as the two studied here. While future epidemiological studies will be required to determine the extent to which this response may protect against endemic HCoV infection, our results suggest that any such protection may be of limited duration, at least to the extent that it is antibody mediated. Nevertheless, it may be possible to exploit the cross-reactivity characterized here through rational immunization or therapeutic strategies designed to elicit escape-resistant immunity across the CoV family.

## Methods

### Study subjects

Under an IRB-approved study (WIRB#1299650), 21 healthy participants were recruited from a local research institution and the surrounding community. Subjects donated blood prior to their first dose (baseline) of the Moderna COVID-19 vaccine (mRNA-1273), approximately 8 days following the first dose (“day 8”), just prior to the second dose (“day 28”) and then approximately 140 days from baseline (“day 140”). The characteristics and exact collection timepoints for each donor are listed in Supplemental Table 1.

Under a separate IRB-approved study (IRB#20204, NCT04497779), unvaccinated COVID-19 convalescent individuals were recruited from the participating clinical sites. Participants all experienced mild COVID-19 symptoms and donated blood within 7-142 days of their initial PCR-based diagnosis. Age, gender, diagnosis and sample collection dates are listed in Supplemental Table 2.

### ELISA analysis

Total SARS-CoV-2 Spike-binding or RBD:ACE2 inhibiting antibody levels were quantified in plasma using ELISA kits: SCoV-2 *Detect*™ IgG (InBios) or SARS-CoV-2 Surrogate Virus Neutralization Test (GenScript), respectively, according to the manufacturer’s instructions.

### HV2 library design

The Human Virome version 2 (“HV2”) PepSeq library included 15,000 unique 30mer peptides and was designed to cover the most reactive linear epitopes from 80 virus species commonly known to infect humans, including five species of CoVs (SARS-CoV, HCoV-OC43, HCoV-229E, HCoV-NL63 and HCoV-HKU1). Most of the peptides in HV2 (70%; 10,536/15,000) were selected from our original HV (10,475) or SCV2 (61) libraries (Ladner et al., 2021) based on empirical observations of reactivity in human sera. An additional 3566 peptides (24% of HV2) were selected to ensure that we included homologous regions from all viruses that belong to the same genus (see Supplemental Methods for details). The additional peptides were used 1) to increase coverage of underrepresented viruses by including publicly reported linear epitopes obtained from IEDB (404 peptides), 2) to finely map reactive sites through alanine scanning (480 peptides) and 3) for negative controls (14 peptides) (see Supplemental Methods for details).

### PepSeq library synthesis and assay

The HV2 library was encoded as a library of 15,000 DNA oligonucleotides and used to synthesize a corresponding ‘PepSeq’ library of DNA-barcoded peptides for multiplexed analysis of plasma antibodies, as previously described (Ladner et al., 2021). Briefly, the oligonucleotide library was PCR-amplified and then used to generate mRNA in an *in vitro* transcription reaction. The product was ligated to a hairpin oligonucleotide adaptor bearing a puromycin molecule tethered by a PEG spacer and used as a template in an *in vitro* translation reaction. Finally, a reverse transcription reaction, primed by the adaptor hairpin, was used to generate cDNA, and the original mRNA was removed using RNAse. To perform serological assays, the resulting DNA-barcoded peptide library was added to diluted plasma and incubated overnight. The binding reaction was applied to pre-washed protein G-bearing beads, washed, eluted, and indexed using barcoded DNA oligos. Following PCR cleanup, products were pooled, quantified and sequenced using an Illumina NextSeq instrument.

### PepSeq data analysis

PepSeq sequencing data was processed and analyzed as previously described (Ladner et al., 2021) using PepSIRF v1.4.0(Fink et al., 2020), as well as custom scripts (https://github.com/LadnerLab/PepSIRF/tree/master/extensions). First, the reads were demultiplexed and assigned to peptides using the PepSIRF *demux* module, allowing for one mismatch in each index sequence and two mismatches in the variable DNA tag region. The PepSIRF *norm* module was then used to normalize counts to reads per million (RPM). RPM normalized reads from seven buffer-only control samples were subsequently used to create bins for Z-score calculation using the PepSIRF *bin* module. To normalize for different starting peptide abundances within each bin, reads were further normalized by subtracting the average RPM from the buffer only controls (--diff option in *norm* module). Z-scores were calculated using the PepSIRF *zscore* module using the 75% highest density interval within each bin. Finally the PepSIRF *enrich* module was used to calculate peptides that were enriched in each sample using a minimum Z-score threshold of 10.

For epitope level analysis (**Figure 2**), each sample was run in duplicate and epitope reactivity was calculated by taking the peptide with the maximum Z-score in either of the two replicates. If multiple peptides contained the target epitope sequence the peptide with the highest signal was selected.

Reduction in reactivity following peptide depletion was calculated by comparing depleted and non-depleted Z-scores across all HV2 peptides. For each sample, a linear spline was fit to the data and reductions in reactivity were calculated by taking the difference between the depleted Z-score and the spline predicted Z-score for each peptide of interest. Only samples with average peptide Z-scores above 6 prior to depletion were analyzed. If multiple peptides contained the epitope of interest, the peptide with the largest difference was used.

To assess the importance of each residue within 30mer peptides containing the FP and SH epitopes, we compared the Z scores for peptides containing alanine substitutions at each peptide residue to the Z score of the wild-type peptide. We refer to this metric the “residue dependency index” and it was calculated as follows: log_2_(Z_WT_ + 3) - log_2_(Z_ala_ + 3), where Z_WT_ and Z_ala_ are the Z scores for the wild-type and alanine substituted peptides, respectively. To look at changes over time in antibody binding profiles, we compared estimates of this residue dependency index between days 28 and 140 post vaccination. To evaluate the significance of changes in the residue dependency index within the SARS-CoV-2 peptides, we also calculated the same metric for residues contained within non-CoV peptides for which alanine mutants were also included in HV2 (i.e. negative controls). In total, we considered 11 negative control peptides each designed from a different common human-infecting virus. Subject/peptide combinations were included as negative controls in the analysis if the wild-type peptide had a Z score ≥ 10 at both days 28 and 140 post-vaccination (i.e., strong reactivity at both time points) and if the Z score fold change between these time points was less than 1.5 (i.e., no substantial change in reactivity between time points).

### Species-specific antibody depletion

In selected cases, species-specific subsets of antibodies were depleted from plasma samples using bead-bound peptides prior to the PepSeq assay. Pairs of chemically synthesized N-terminally biotinylated 20-mer FP + SH peptides (Sigma) with sequences from SARS-CoV-2 (PSKRSFIEDLLFNKVTLADA + LQPELDSFKEELDKYFKNHT) or HCoV-OC43 (ASSRSAIEDLLFDKVKLSDV + SIPNLPDFKEELDQWFKNQT) were pooled at an equimolar ratio (100 ug/mL), added to 180uL of pre-washed Streptavidin beads (Thermo) and incubated at room temperature for 15 minutes on a rotator. Peptide-coated beads were then washed 3 times and re-suspended in superblock (Thermo). Next, 60uL of serum/plasma was added to 60uL of peptide-coated beads and incubated for 15 minutes on a rotator. Serum was removed from the beads and re-applied to a fresh peptide-bearing bead aliquot and repeated, for a total of 3 depletion cycles, prior to use in the PepSeq assay.

### Full-length Spike protein reactivity (MagPix)

The SARS-CoV-2 spike protein (Isolate WA1) was kindly provided by InBios International, Inc. (Seattle Washington). The HCOV-OC43 spike (S1+S2 ECD His Tag) protein was purchased from Sino Biological. Nucleocapsid proteins (used as controls) were synthesized in-house. Purified His-tagged proteins from SARS-CoV-2 (spike and nucleocapsid), HCoV-NL63 (nucleocapsid), and HCov-OC43 (spike) viruses along with immunoglobulin G (ThermoFisher Scientific, Waltham, MA) and R-Phycoerythrin (AnaSpec, Fremont, CA) were covalently conjugated to different fluorescently labeled MagPlex® microspheres using the two-step carbodiimide coupling chemistry at pH 5-6 using Sulfo-NHS (N-hydroxysulfosuccinimide) and EDC (1-ethyl-3-(3-dimethylaminopropyl) carbodiimide hydrochloride) (Luminex xMAP® Cookbook, 4th Ed.). The Purified His-tagged proteins were conjugated with 5 μg protein per one million beads ratio. The immunoglobulins and R-Phycoerythrin conjugated with 0.5 μg protein per one million beads ratio. After the conjugation, the conjugated microspheres were stored in PBS-TBN (0.05% Tween, 1.0% BSA, 0.1% Sodium Azide) at 4 °C with a light block. His-tagged protein bead conjugation was confirmed using monoclonal anti-6-histidine antibody (Abcam, Boston, MA) conjugated to biotin and Streptavidin, R-Phycoerythrin Conjugate (SAPE, Life technologies). The antibody conjugation was confirmed using goat anti-human IgG biotin (Abcam, Boston, MA) and SAPE.

Protein conjugated beads were diluted in 1x Blocker BSA solution (Fisher Scientific, Hampton, NH) and mixed at a ratio of 1,000 beads per bead region per assay. Beads were washed two times with wash buffer (11.9mM Phophate, pH 7.4, 137mM NaCl, 2.7mM KCl, and 0.05% Tween 20) and the wash buffer was removed after beads were bound to a plate magnet. Serum was diluted 500 or 2,500-fold in 1x Block BSA solution and 100μl diluted serum was added to the beads. Serum and beads were incubated for 1 hour with shaking at room temperature. The beads were washed 3 times with wash buffer and 100 ul of 2 mg/ml Goat anti human IgG (Abcam, Boston, MA) biotin conjugated secondary antibody diluted in 1x Block BSA were added to the beads. The secondary antibody and beads were incubated for 1 hour with shaking at room temperature and washed 3 times with wash buffer after incubation. Beads were then incubated with 4 mg/ml SAPE (Life Technologies) diluted in 1x Blocker BSA for 0.5 hours with shaking at room temperature and washed 3 times with wash buffer after incubation. Beads were suspended in 1x Blocker BSA and the signal intensities of different fluorescently labeled MagPlex® microspheres were read on a MAGPIX^®^ system (Luminex, Austin, TX) and the median fluorescence intensity (MFI) units per bead region was calculated using xPONENT^®^ software (Luminex). MFI intensities were corrected based on sample dilution to ensure linearity of the assay.

### Statistical analysis

For epitope discovery, PepSeq Z-scores (calculated as described above) from CoV peptides were log2 transformed and compared across the 4 timepoints, matched by subject using repeated-measures ANOVA. P-values were corrected for the False Discovery Rate across the 421 CoV peptides, and a corrected alpha threshold value of 0.05 was used. To evaluate the responses at defined epitope regions, Paired T-tests were used to compare log2 transformed Z-scores for focal peptides between each post-vaccine timepoint and baseline, for homologous peptides within timepoints, or to compare the MagPix intensities from full-length SARS-CoV-2 or HCoV-OC43 Spike proteins between each post-vaccine timepoint and baseline. For the cross-sectional convalescent cohort, an unpaired T-test was used to compare the response maturity ratio for the 2 epitopes of interest between donor groups dichotomized by time since COVID diagnosis.

## Data Availability

All data produced in the present study are available upon reasonable request to the authors

## Acknowledgments

We are grateful to the vaccinated and convalescent blood donors, Carmel Plude for phlebotomy, Danielle Metz and Bethine Moore for clinical research co-ordination, and Haley Nunnallay for technical assistance in generating the MagPix data. We also thank Dr Daniela A Bota for her assistance in recruiting convalescent donors. This work was supported by the NIAID (U24AI152172, U24AI152172-01S1, U24AI152172-IOF, PI: J.A.A.), the California Institute for Regenerative Medicine (CLIN2COVID19-11775, PI: J.A.Z), the State of Arizona Technology and Research Initiative Fund (TRIF, administered by the Arizona Board of Regents, through Northern Arizona University), the Flinn Foundation, and The Cowden Endowment for Microbiology. The contents of this publication are solely the responsibility of the authors and do not necessarily represent the official views of CIRM or any other agency of the State of California.

## Supplemental Material

**Supplemental Figure 1:**
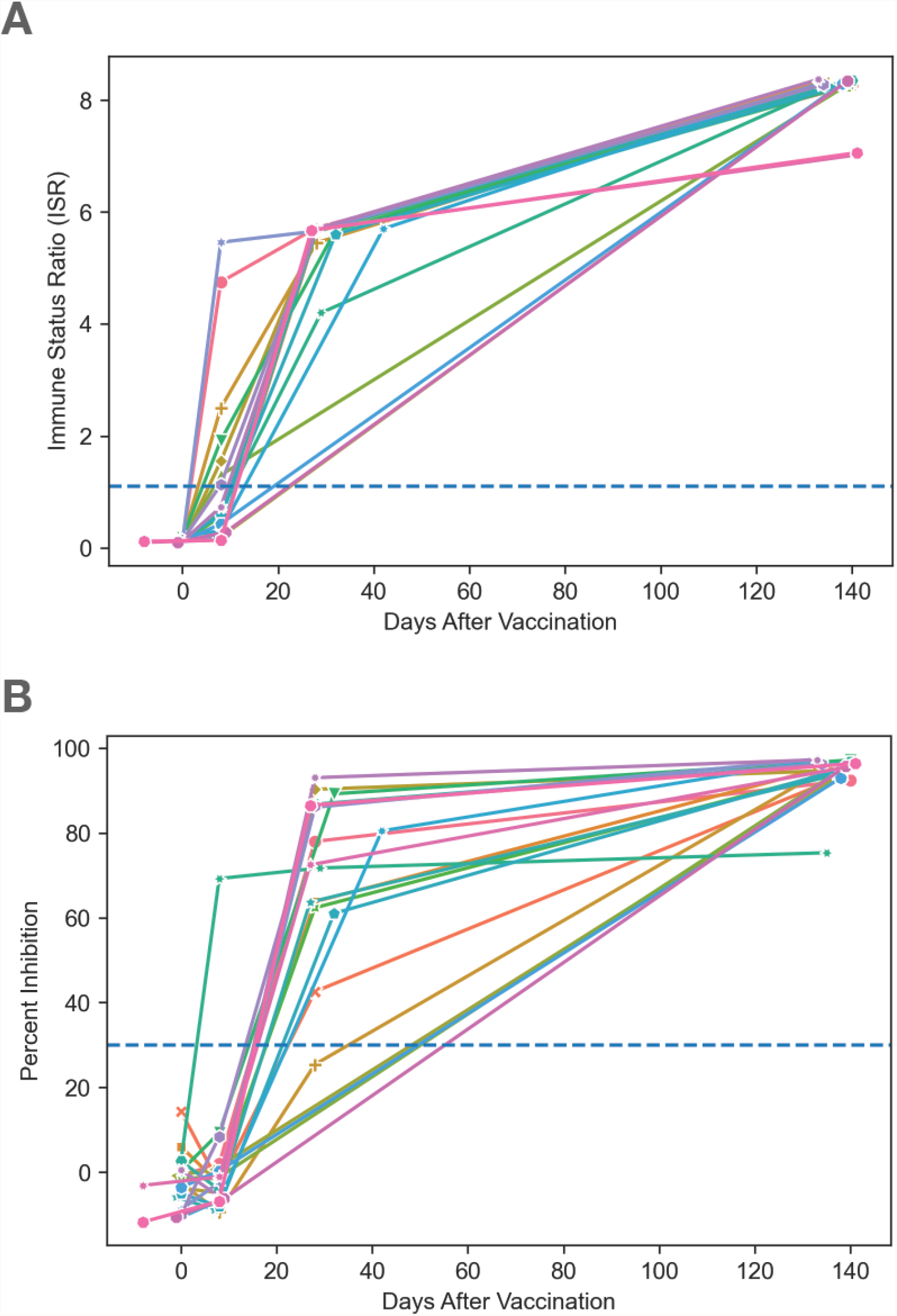
Vaccine-induced increases in SARS-CoV-2 Spike-binding and RBD-inhibiting antibodies over time across the cohort. **A**. Magnitude of plasma IgG binding to SARS-CoV-2 Spike over time in vaccinated patients, measured by ELISA (InBios). Positivity threshold (ISR = 1.1) is indicated as dashed blue line. **B**. Magnitude of plasma inhibition of SARS-CoV-2 RBD binding to ACE2 over time in vaccinated patients, measured by competition ELISA (GenScript). Positivity threshold (30% inhibition) is indicated as dashed blue line.

**Supplemental Table 1.** Vaccinated Participant Biodata.

| Sex | Age range | Period between doses (days) | Collection timepoints<br>(days from first vaccine dose) |  |  |  |
| --- | --- | --- | --- | --- | --- | --- |
|  |  |  | ~Day 0 | ~Day 8 | ~Day 28 | ~Day 140 |
| Female | 30-39 | 28 | 0 | 8 | 28 | 140 |
| Female | 30-39 | 28 | 0 | 8 | 28 | 138 |
| Male | 30-39 | 28 | 0 | 8 | 28 | 133 |
| Male | 40-49 | 28 | 0 | 8 | 28 | 135 |
| Female | 30-39 | 28 | 0 | 8 | 28 | 138 |
| Male | 50-59 | 32 | 1 | 7 | 28 | 138 |
| Female | 18-29 | 28 | 0 | 8 | NC | 139 |
| Male | 40-49 | 28 | 0 | 8 | 28 | 140 |
| Male | 18-29 | 32 | 0 | 8 | 32 | 140 |
| Male | 60+ | 29 | 0 | 8 | 29 | 135 |
| Female | 18-29 | 28 | 0 | 8 | 28 | 140 |
| Female | 18-29 | 27 | 1 | 7 | 27 | 137 |
| Male | 18-29 | 32 | 0 | 8 | 32 | 138 |
| Female | 40-49 | 45 | 0 | 8 | 42 | 134 |
| Male | 18-29 | 28 | 0 | 8 | NC | 138 |
| Female | 30-39 | 28 | 0 | 8 | 28 | 133 |
| Female | 50-59 | 28 | 2 | 8 | 28 | 134 |
| Female | 30-39 | 28 | 0 | 8 | 28 | 133 |
| Female | 60+ | 28 | 1 | 10 | 25 | NC |
| Female | 60+ | 28 | 8 | 8 | 27 | 141 |
| Male | 60+ | 28 | 8 | 8 | 27 | 141 |

NC= Not Collected, \*Prior to first vaccine
All timepoints are expressed in days relative to the day on which the first vaccine dose occurred. Day 28 collection occurred before the second vaccine dose.

**Supplemental Table 2.** Convalescent Participant Biodata.

| Sex | Age range | Collection timepoint<br>(days from positive<br>diagnosis) |
| --- | --- | --- |
| Male | 50-59 | 38 |
| Female | 30-39 | 121 |
| Female | 40-49 | 85 |
| Female | 40-49 | 68 |
| Female | 30-39 | 69 |
| Female | 30-39 | 53 |
| Female | 50-59 | 94 |
| Female | 18-29 | 29 |
| Female | 40-49 | 127 |
| Female | 30-39 | 81 |
| Female | 40-49 | 78 |
| Male | 18-29 | 44 |
| Female | 60+ | 135 |
| Female | 50-59 | 118 |
| Male | 30-39 | 59 |
| Female | 18-29 | 50 |
| Female | 30-39 | 40 |
| Female | 40-49 | 66 |
| Female | 30-39 | 119 |
| Female | 18-29 | 70 |
| Female | 18-29 | 99 |
| Male | 40-49 | 67 |
| Female | 50-59 | 74 |
| Female | 18-29 | 79 |
| Female | 50-59 | 111 |
| Male | 30-39 | 49 |
| Female | 30-39 | 92 |
| Male | 60+ | 142 |
| Male | 40-49 | 69 |
| Female | 50-59 | 50 |
| Male | 30-39 | 65 |
| Female | 50-59 | 20 |
| Female | 30-39 | 34 |
| Male | 50-59 | 20 |
| Female | 60+ | 30 |
| Female | 60+ | 28 |
| Female | 18-29 | 24 |
| Female | 50-59 | 17 |
| Female | 30-39 | 28 |
| Female | 50-59 | 25 |
| Female | 50-59 | 25 |
| Female | 18-29 | 30 |
| Female | 30-39 | 24 |
| Female | 50-59 | 21 |
| Male | 30-39 | 31 |

Positive diagnosis determined by PCR. Collection time point is expressed in days relative to the day on which the patient tested PCR positive for COVID-19.

**Supplemental Table 3.**
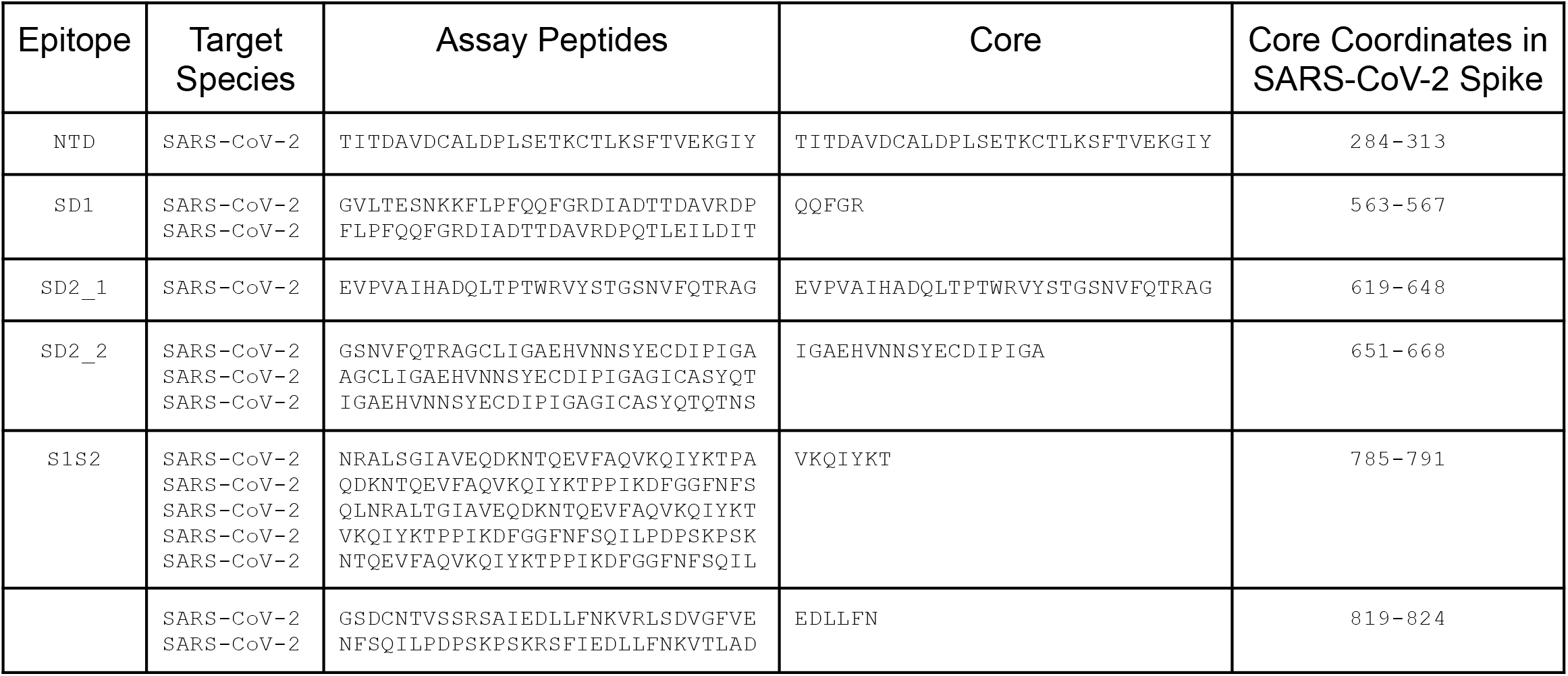
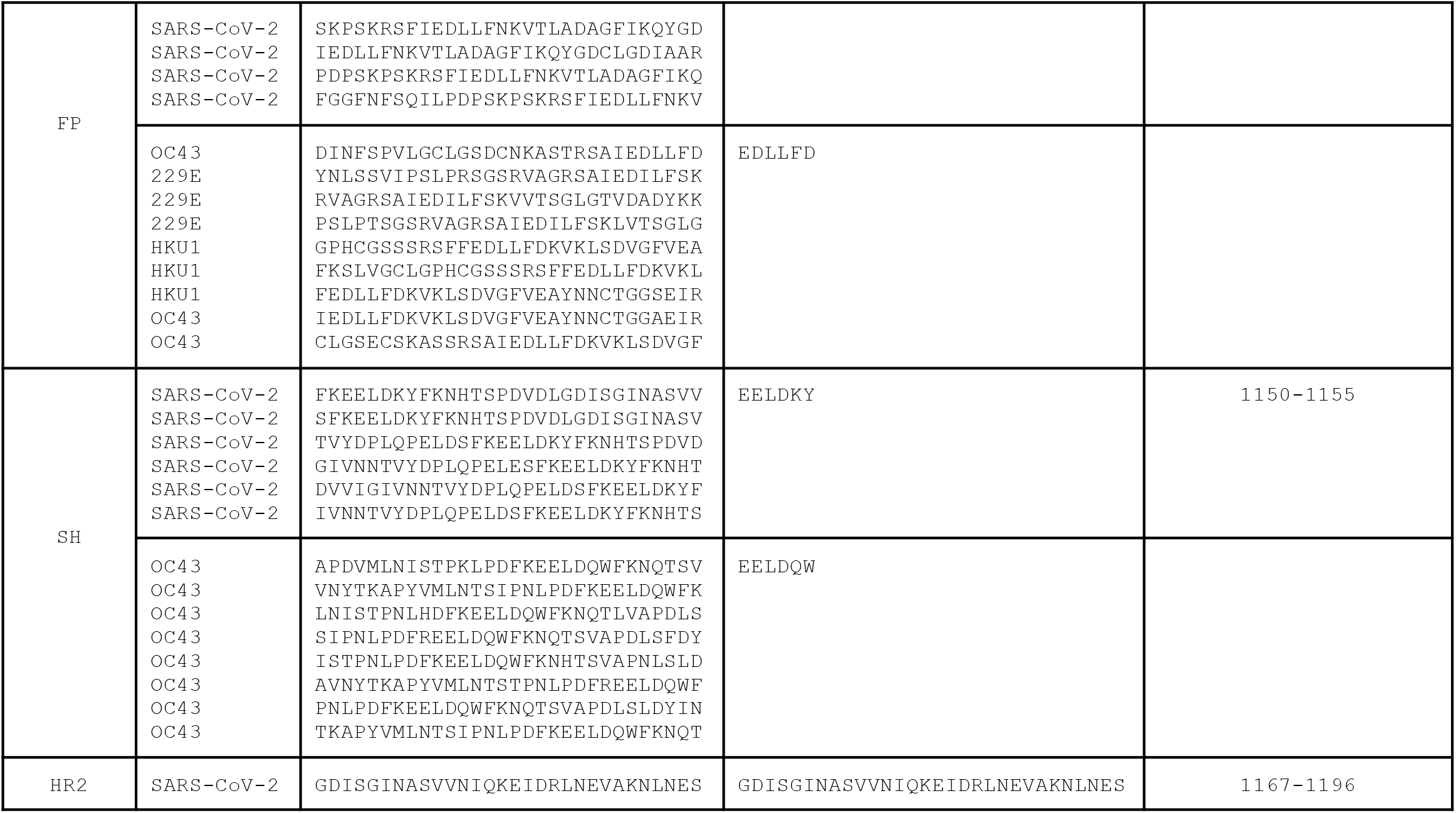
PepSeq Coronavirus Peptides.

## Supplemental Text

### HV2 Library Design

To increase coverage of underrepresented viruses in HV2, we included publicly reported linear epitopes present in IEDB (https://www.iedb.org/). Specifically, we downloaded 913 linear B cell epitopes covering 17 virus species: Alphapapillomavirus 7, Alphapapillomavirus 9, Chikungunya virus, Human gammaherpesvirus 8, Human immunodeficiency virus 1, Human mastadenovirus A, Human polyomavirus 1, Human polyomavirus 2, Japanese encephalitis virus, Lymphocytic choriomeningitis mammarenavirus, Mumps rubulavirus, Orthohepevirus A, Primate T-lymphotropic virus 1, Rabies lyssavirus, Rubella virus, West Nile virus, and Zika virus. All IEDB epitopes were ≤48 aa long. For each epitope >30 aa, two peptides were designed, one starting at the N terminus and one ending at the C terminus. For each epitope ≤30 aa, a single peptide was designed, with extra residues obtained, when necessary, from the longer protein sequence to which the epitope was linked. The final HV2 design included 404 peptides designed with this approach.

One of our goals with the design of the HV2 PepSeq library was to ensure the inclusion of homologous peptides from closely related viruses. Specifically, we focused on 12 genera, each of which contained ≥2 virus species in our design: Alphacoronavirus, Alphapapillomavirus, Betacoronavirus, Betapapillomavirus, Betapolyomavirus, Enterovirus, Flavivirus, Mastadenovirus, Roseolovirus, Rotavirus, Rubulavirus, and Simplexvirus. For each genus, we generated protein-level alignments with one representative from each species. Then, for each HV2 peptide selected from HV1, SCV2 or IEDB, we designed peptides covering the homologous region from all congeners. However, these new peptides were only included in the final HV2 design if they were distinct from all existing design peptides (<70% of 5mers shared). The final HV2 design included 3566 peptides designed with this approach.

To finely map antibody binding sites within a subset of peptides, we also include 30 derivative peptides, one with an alanine (or glycine, if alanine was the wild-type amino acid) in place of the wild-type residue at each residue (“alanine scans”). Specifically, we included these alanine scans for 16 unique peptides, including peptides covering the SARS-CoV-2 FP and SH epitopes. The final HV2 design included 480 peptides designed with this approach.

Finally, we included 14 negative control peptides derived from an assortment of eukaryotic proteins of exotic species (e.g., coelacanth, coral).

